# Clinical Holds in Early Oncology Drug Development

**DOI:** 10.1101/2022.06.03.22275971

**Authors:** Alexandra Snyder, Dinesh De Alwis, Asthika Goonewardene, Priti S. Hegde

## Abstract

Clinical holds in oncology are necessary to safeguard patients. Researchers are alerted to holds by news sources, but a systematic evaluation of clinical holds in early oncology drug development is lacking. Analysis of publicly disclosed clinical holds in oncology from 2016-2021 identified 39 holds. The majority (n=29) were for toxicity-related reasons, with fewer for chemistry, manufacturing and controls (CMC) (n=7) or other reasons (n=3). Toxicity-related holds took a median of 74 days till resolution, whereas chemistry, manufacturing and controls (CMC)-related holds took a median of 108 days to resolve. Acknowledging the limited sample size and scope of stock market conditions, toxicity-related clinical holds impacted the market value of small/medium sized biotechs by a median of -15%, which is far more than large biopharma (median 0%). These data suggest that toxicity-related clinical holds are common in early oncology and can have a more detrimental impact on small/medium sized biotech sponsors, the latter of which could have substantial financial repercussions, particularly in “biotech bear markets” where public investing in the sector has fallen out of favor (as in 2021-2022).

## Text

Development of novel oncology drugs and platforms now occurs primarily in the context of smaller biotechnology companies. In the years 2020-21, 36% of oncology drug approvals by the US FDA were of drugs developed by a biotech company and 64% by big pharma (Supplementary Table S1). Importantly, a large majority of approvals for big pharma constituted label expansions, whereas 100% of approvals for small biotech were related to novel therapies or platforms^1^. Indeed, approximately 80% of the overall industry pipeline comes from emerging biopharma companies^2^. Of the 112 oncology drug approvals in 2020-2021, 40 were of drugs developed by small biotechs. Of these 40, based on company disclosures, 9 were put on partial or full clinical hold at some point in their development. Such holds have at times led to a dramatic drop in public interest in a biotech, despite evidence of efficacy (Fig. 1A).

**Figure 1A.**
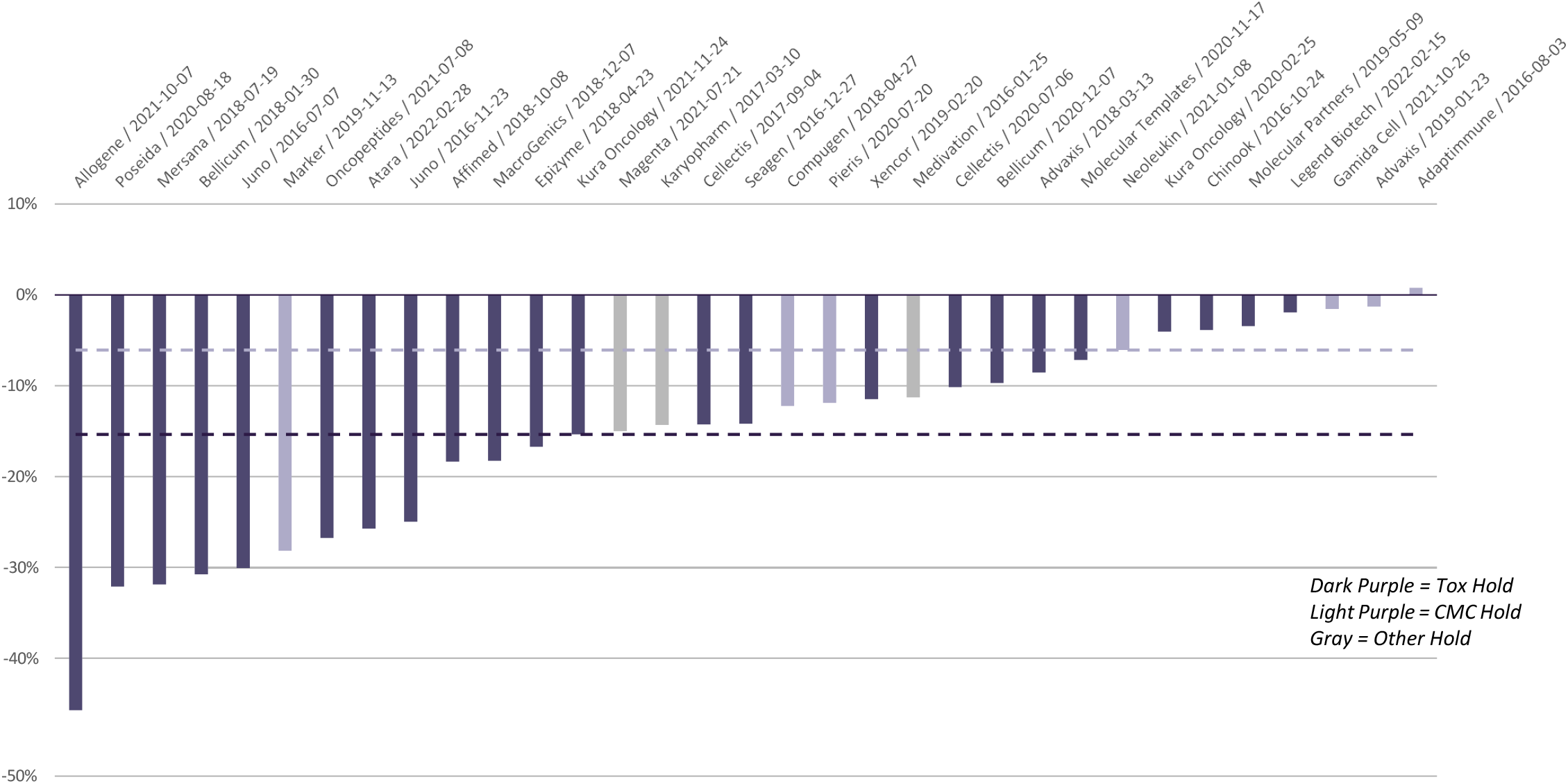
Share Price Reaction to Clinical Hold Announcement - SMID Biotech This chart represents the share price movement on the day of or day after a clinical hold was first reported, against the prior 3 day rolling average. The X axis indicates the company and the date the hold was announced. Dark purple bars represent share price reactions for clinical holds due to toxicity, light purple bars represent clinical holds due to CMC issues, and gray bars represent clinical holds due to other reasons (PK/PD, trial disclosures, etc.). Dotted lines represent median share price reaction to clinical holds due to toxicity (dark purple) or CMC (light purple). In this chart, only SMID-cap biotech companies’ share price reactions and medians are represented. SMID=small and medium cap biotechs, here defined as value less than $10B. Large cap defined as value greater than or equal to $10 billion.

Chimeric antigen receptor (CAR) therapies provide an example of platform development that has benefited from time spent in understanding clinical safety, including revisions caused by clinical holds. CARs outfit T cells and other immune cell types with receptors that can attack cancer and have now become standard of care for the treatment of some patients with lymphoma and multiple myeloma. However, their development path has featured pauses and redirections, as these drugs can lead to substantial systemic inflammation, including cytokine release syndrome (CRS) and immune effector cell-associated neurotoxicity syndrome (ICANS). A study of a CD19 CAR-T therapy, JCAR015, was put on hold in 2016 due to 3 patient deaths out of 20 accrued patients. Initially, fludarabine, a drug used to condition the body prior to the experimental therapy, was faulted. However, after an additional 2 patients died, the trial was put on hold indefinitely and the sponsor, Juno, shifted development to JCAR017, an optimized CAR-T drug that also targeted CD19 but with different viral vector, binding domain, costimulatory domain and T-cell selection process. Juno lost $1.36B in market cap when they announced the first clinical hold in July 2016, and $1.39B when they announced the second hold in November 2016 (Supplementary Fig. S1), although it bears noting that other fluctuations occurred in the stock price outside of these events. The optimized agent, JCAR017, was ultimately approved by the FDA for treatment of lymphomas^3^ ; this success, among other variables, has spawned vigorous development and optimization in the cell therapy space.

To identify larger trends in the field, as there is no repository of clinical holds in early oncology, we surveyed press releases and public investor call transcripts with the words “clinical hold” available on Bloomberg from 2016 to 2021 for US-based companies (Table 1). 39 oncology-related holds occurred during this period. Of these, among the 29 toxicity-related holds, 10 were for cell therapies, 6 for T-cell engagers (TCE), 5 for antibodies in immuno-oncology, 4 for antibody-drug conjugates and 4 for small molecules. 18 were resolved, 6 were terminated and 5 were ongoing as of this writing. Seven holds were for reasons related to chemistry, manufacturing and controls (CMC): 3 for cell therapies, 1for TCE and 3 for IO, all of which were resolved. Three holds were for other reasons. Toxicity-related holds took a median of 74 days till resolution irrespective of the platform (with immuno-oncology (IO) agents an outlier at 100 days median) (Table 1B), whereas CMC-related holds took a median of 108 days to resolve.

**Table 1A.**
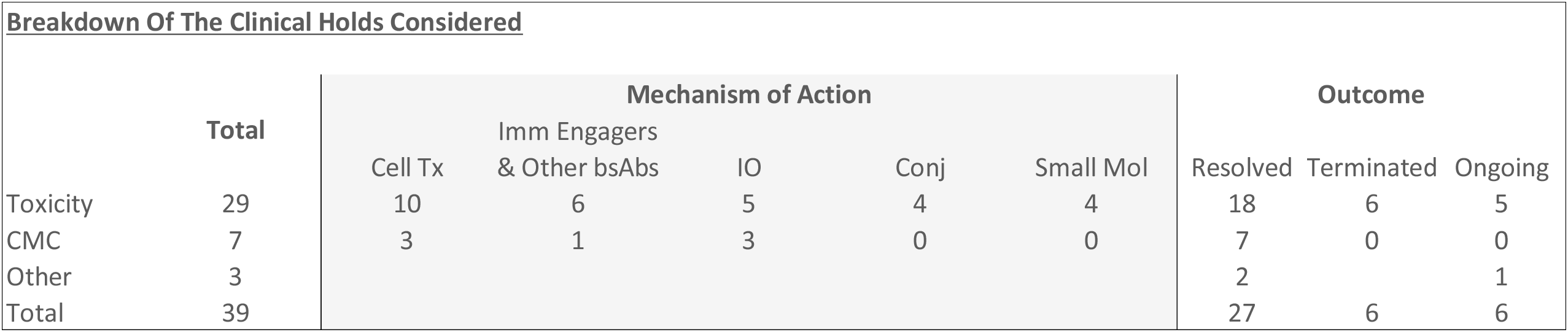
Mechanisms and outcomes of clinical holds in oncology, 2016-2021. CMC = chemistry, manufacturing and controls.

| <u>Breakdown Of The Clinical Holds Considered</u> |  |  |  |  |  |  |  |  |  |
| --- | --- | --- | --- | --- | --- | --- | --- | --- | --- |
|  | Total | Mechanism of Action |  |  |  |  | Outcome |  |  |
|  |  | Cell Tx | Imm Engagers<br>& Other bsAbs | IO | Conj | Small Mol | Resolved | Terminated | Ongoing |
| Toxicity | 29 | 10 | 6 | 5 | 4 | 4 | 18 | 6 | 5 |
| CMC | 7 | 3 | 1 | 3 | 0 | 0 | 7 | 0 | 0 |
| Other | 3 |  |  |  |  |  | 2 |  | 1 |
| Total | 39 |  |  |  |  |  | 27 | 6 | 6 |

**Table 1B.** Median time to resolution of clinical holds, 2016-2021.

| <u>Time To Resolve Clinical Holds (Days)</u> |  |
| --- | --- |
| **Reason for Hold: Toxicity or Patient Death** | **74** |
| *Cell Therapy* | 71 |
| *Immune Engagers & Other Bispecific Antibodies* | 69 |
| *Immuno-Oncology* | 100 |
| *Conjugates* | 65 |
| **Reason for Hold: CMC** | **108** |
| *Cell Therapy* | 98 |

Acknowledging the limited sample size, clinical holds impacted the market value of small/medium sized biotechs more than large biopharma (Table 1C). The rebound after hold removal was greater for CMC-related holds than safety-related holds (Fig. 1B). We hypothesize that this contrast may be because, despite hold resolution, toxicities become part of the risk:benefit profile of the agent, impacting the drug profile beyond the hold.

**Table 1C.** Median share price moves relative to hold announcement, 2016-2021.

| **<u>Median Share Price Moves (On Announcement vs Prior 3 Day Average)</u>** |  |  |  |
| --- | --- | --- | --- |
| Clinical Hold Announced | All companies | SMID-cap | Large-cap |
| *Reason for Hold: Toxicity or Patient Death* | **-11%** | -15% | 0% |
| *Reason for Hold: CMC* | **-6%** | -6% |  |
| Clinical Hold Successfully Resolved |  |  |  |
| *Reason for Hold: Toxicity or Patient Death* | **-1%** | -1% | -1% |
| *Reason for Hold: CMC* | **4%** | 4% |  |
| Clinical Hold Leads To Asset Termination |  |  |  |
| *Reason for Hold: Toxicity or Patient Death* | **-8%** | -8% |  |

**Figure 1B.**
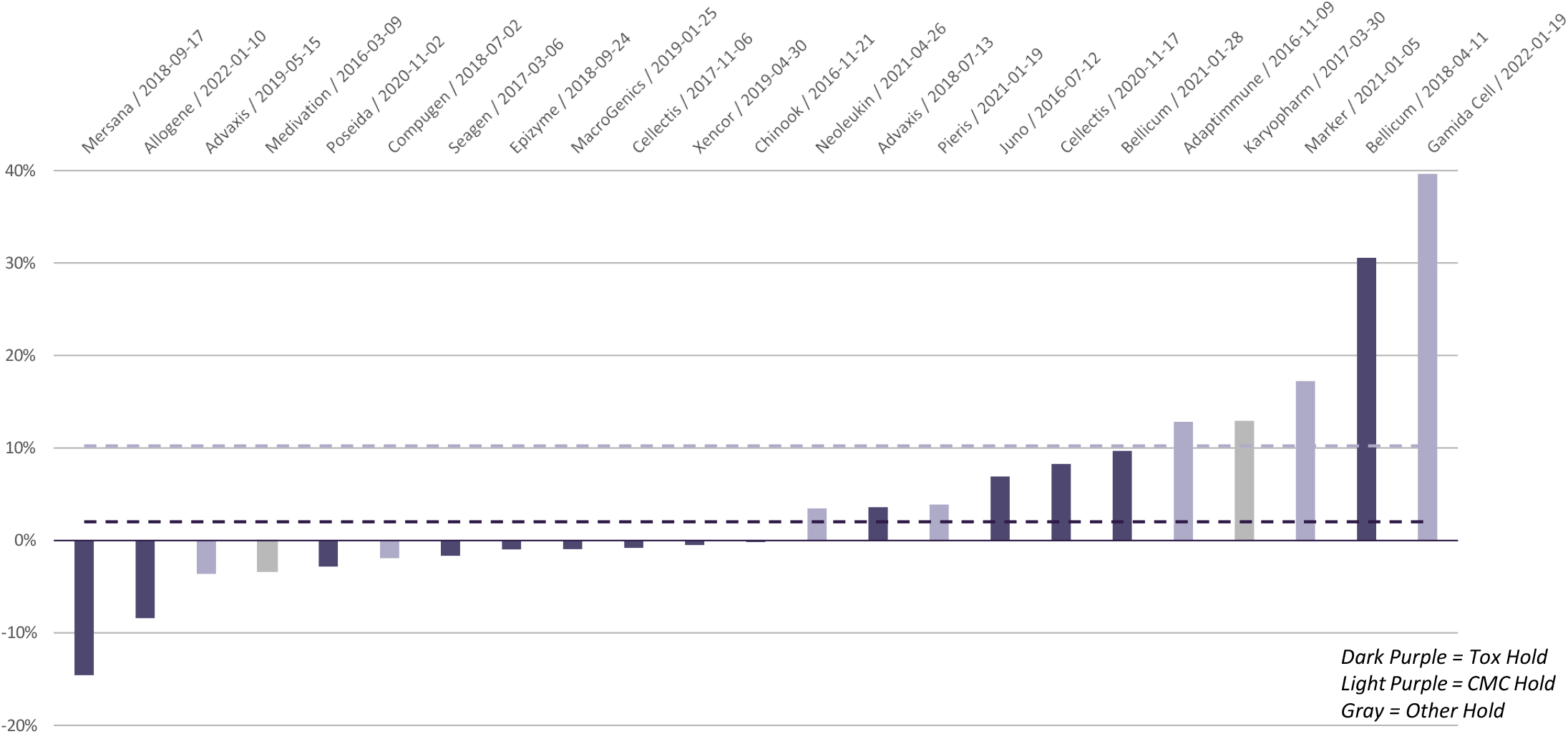
Share Price Reaction to Clinical Hold Resolution - SMID Biotech This chart represents the share price movement on the day of or day after a clinical hold was reported as successfully resolved, against the prior 3 day rolling average. The X axis indicates the company and the date the resolution was announced. Dark purple bars represent share price reactions for clinical holds due to toxicity, light purple bars represent clinical holds due to CMC issues, and gray bars represent clinical holds due to other reasons (PK/PD, trial disclosures, etc.). Dotted lines represent median share price reaction to resolving clinical holds due to toxicity (dark purple) or CMC (light purple). In this chart, only SMID-cap biotech companies’ share price reactions and medians are represented.

### Some limitations of our methodology include

- We were only able to capture clinical holds that were publicly disclosed in a press release or on an investor call; a clinical hold may not be material enough for a large biopharma company to issue a press release, so we may be undercounting clinical holds experienced by larger companies.
- We were only able to capture companies that continued to exist; therefore, our survey likely underestimates the impact of clinical holds on the fate of biotech companies.
- Our analysis of share price reactions accounts for the absolute change in share price, but does not account for the relative movement versus the broader stock market or relevant indices, nor does it account for investor sentiment that is already baked into the share price. Our analysis of events was restricted to 2016-2021, and this represents a period of more favorable market conditions for biotech investing. In more recent times (2021-to date), biotech has been in a “bear market” where long term public market investing has fallen out of favor. In these conditions, negative share price movements due to clinical holds can be exacerbated.

Technological breakthroughs paired with ongoing clinical need make oncology a fertile space for drug development. Such development is attended by myriad risks, including to the safety of patients treated on clinical trials, and requires the consistent production of high quality drug products. Clinical holds are a necessary part of drug development intended to protect patient safety. Historical examples (described in an upcoming publication by Snyder et al.^4^) demonstrate that careful optimization of oncology drugs can lead to the development of therapies that improve and prolong lives; Project Optimus, an FDA initiative start in 2022 to improve dose-finding in oncology, underscores the need for methodical early drug development. While qualitative in nature, data described here suggest that clinical holds, whether for toxicity or CMC, do not necessarily hinder likelihood of eventual drug approval, and may result in better therapeutics. However, such events can cause a significant drop in investor interest, particularly for small biotechnology companies. While such fluctuations are a natural part of drug development, this analysis suggests that a careful examination of each case may be instructive, informing the field whether actions taken as a result of the hold may in fact lead to an approval-worthy drug profile.

## Data Availability

All data produced in the present work are contained in the manuscript.

## Disclosures

AS is an employee of Two River, Inc and owns shares in Allogene Therapeutics and Merck & Co., Inc., Rahway, NJ, USA.

PSH is an employee of Foundation Medicine, Inc and owns shares in Roche, Bolt Therapeutics and TCR2.

DD is an employee of Merck Sharp & Dohme LLC, a subsidiary of Merck & Co., Inc., Rahway, NJ, USA and owns shares in Merck & Co., Inc., Rahway, NJ, USA.

**Supplementary Figure S1.**
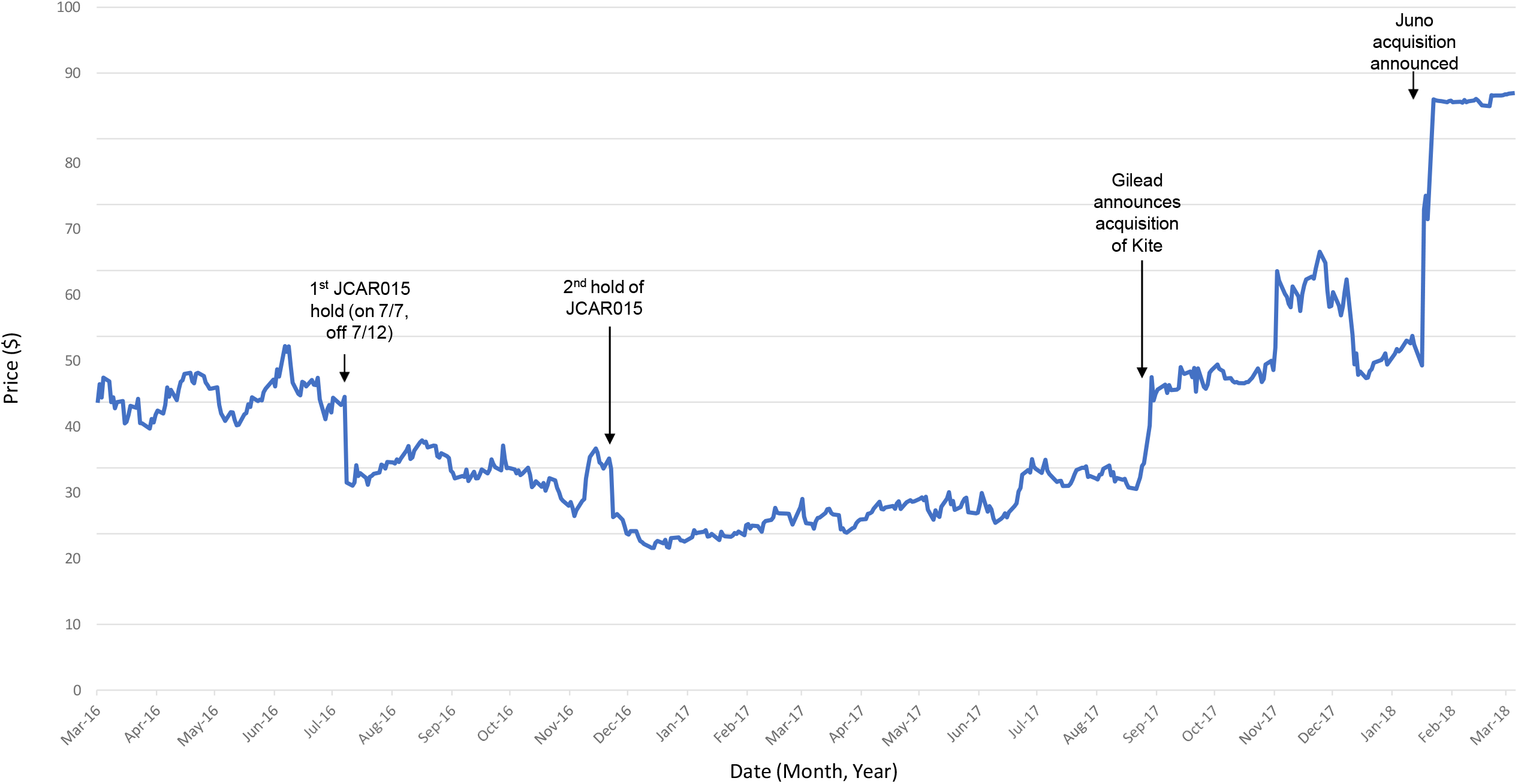
Juno stock price by date and key events, 2016-2018. Source: Bloomberg.

**Supplementary Table S1.**
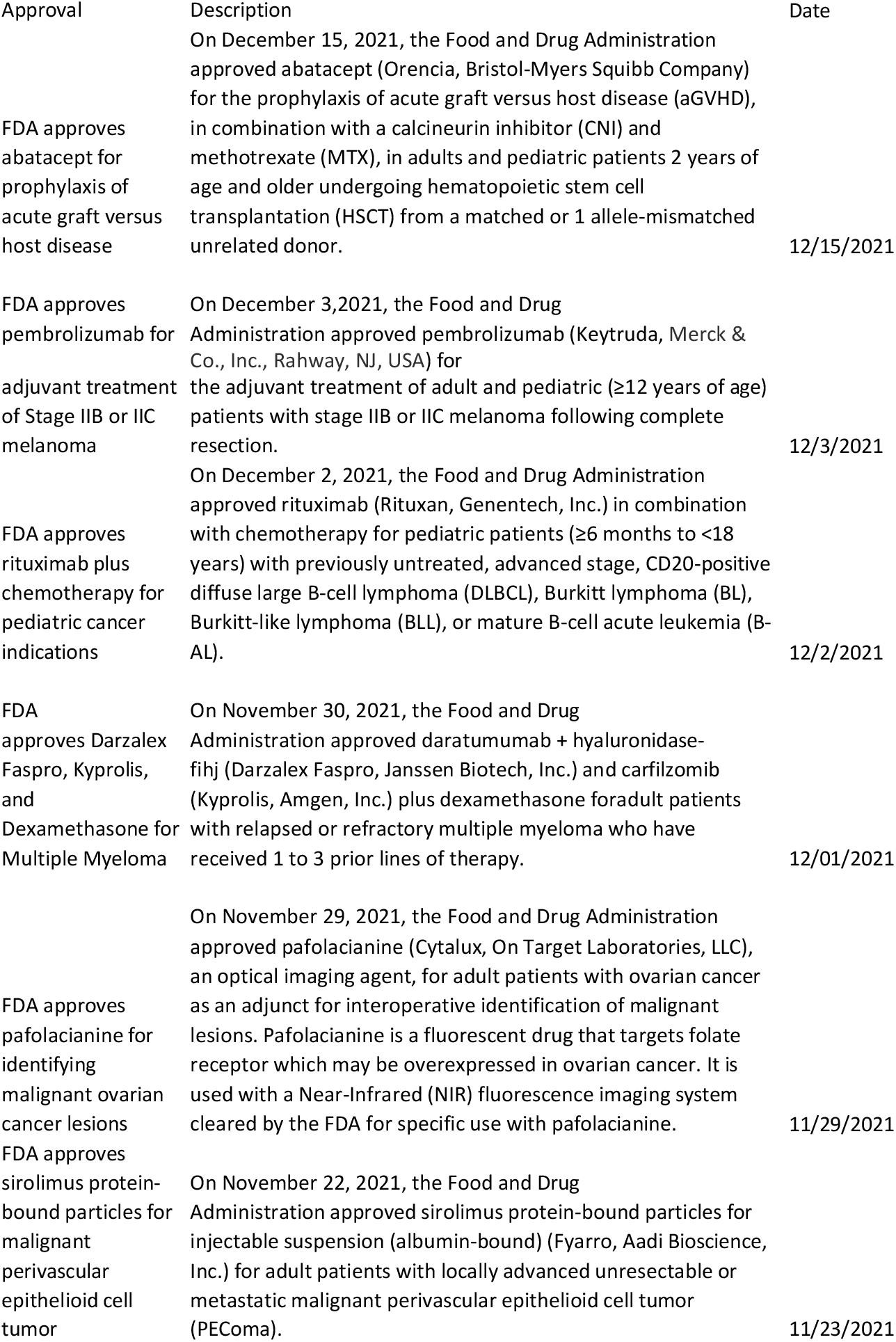
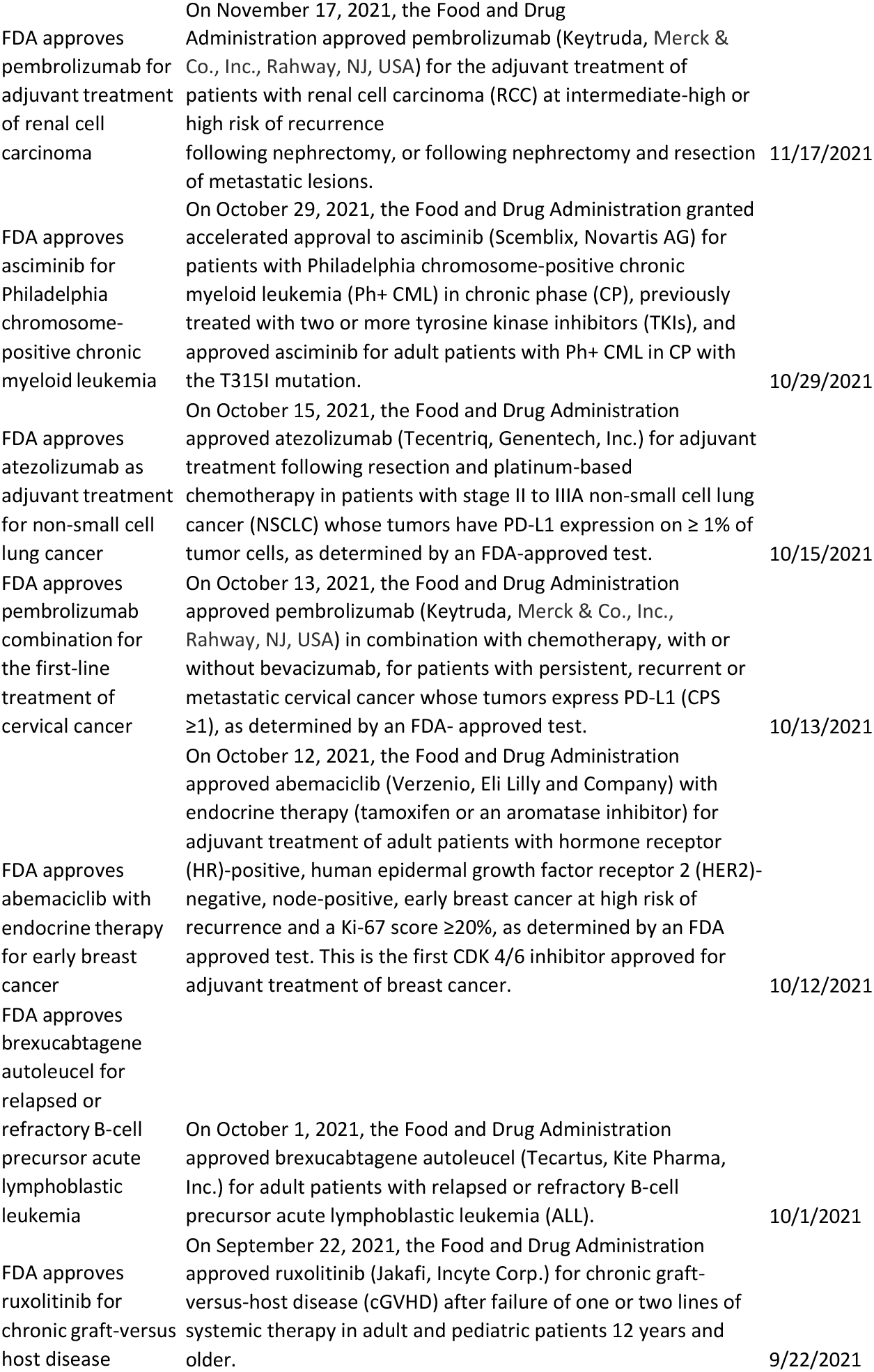
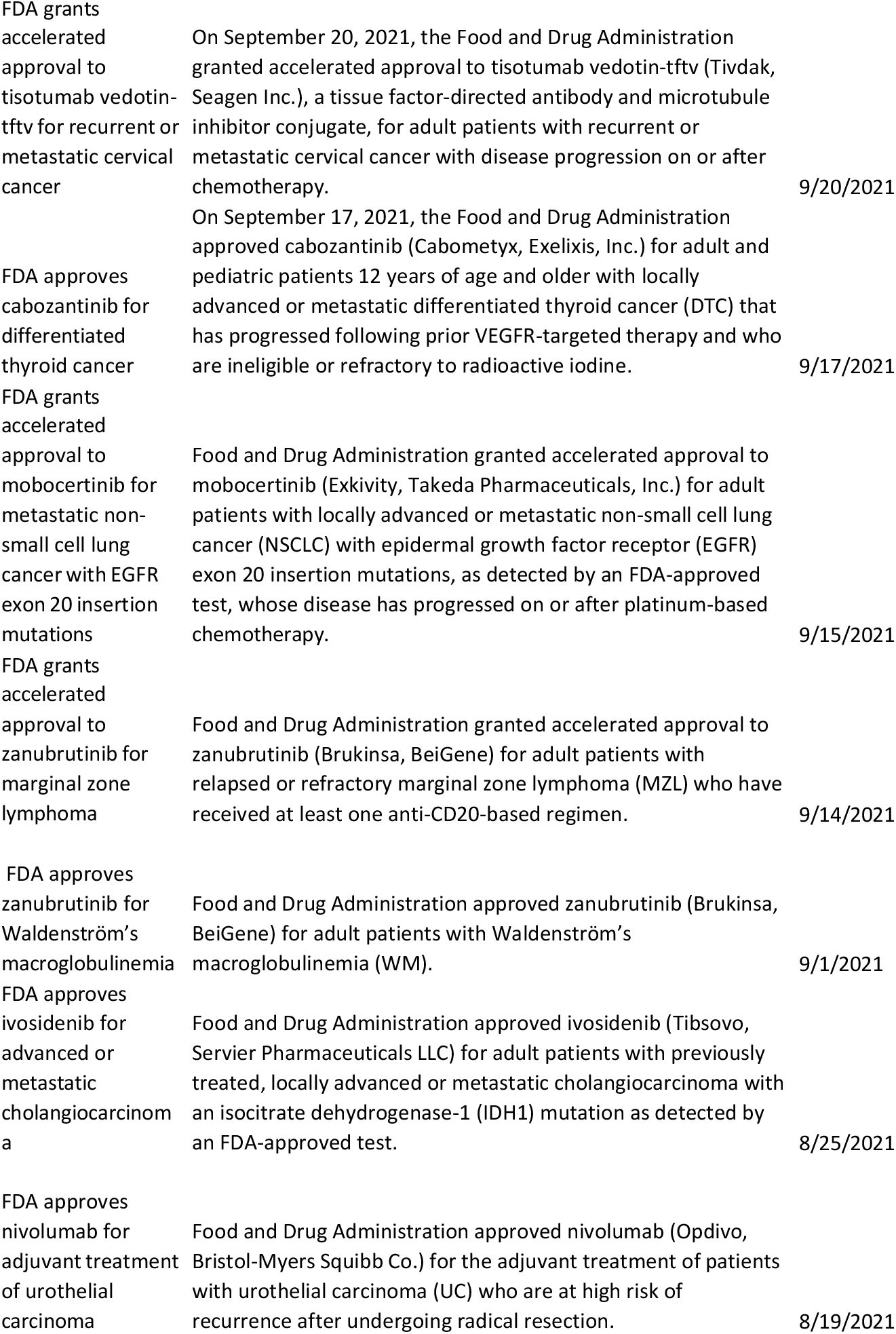
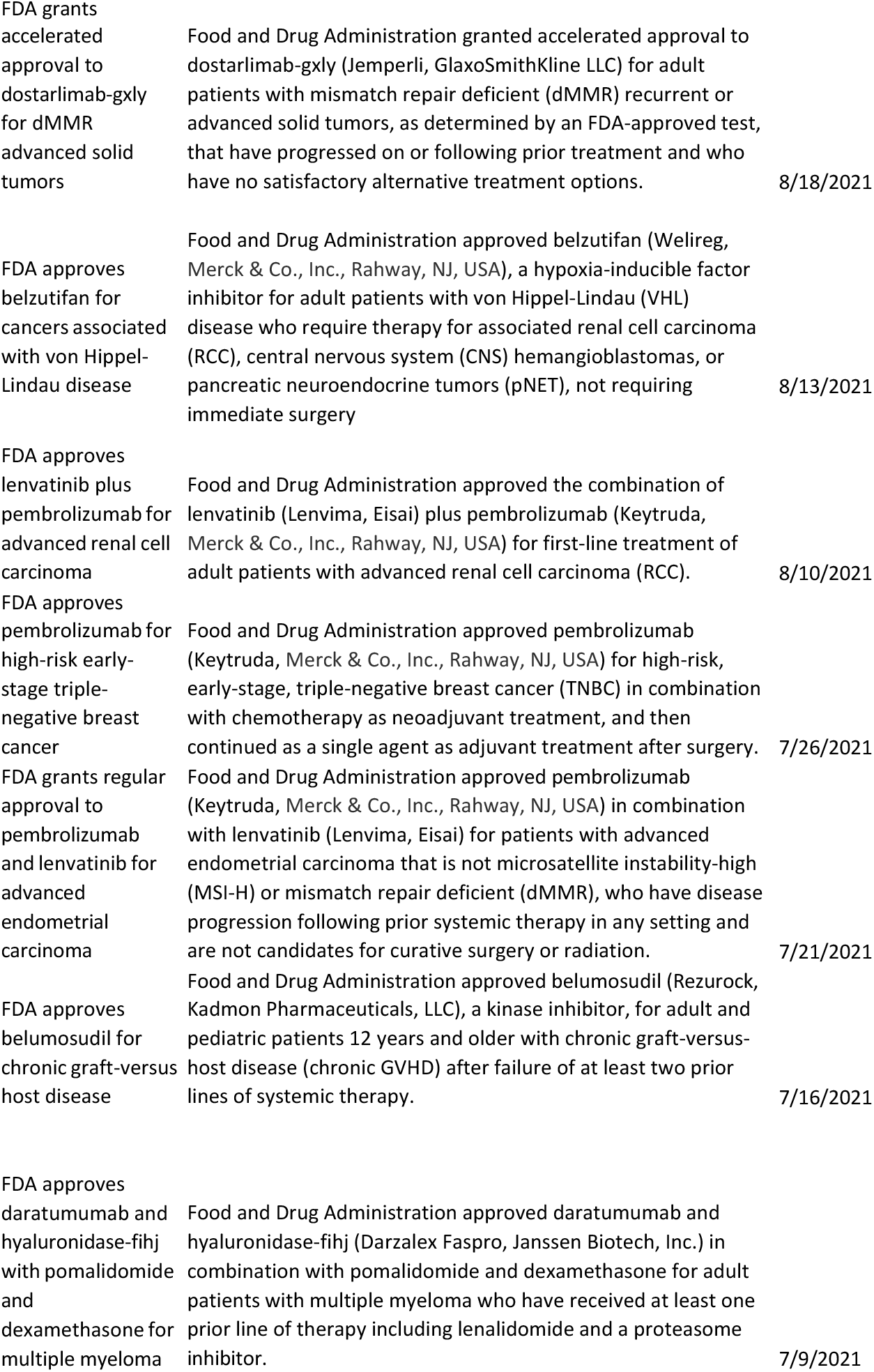
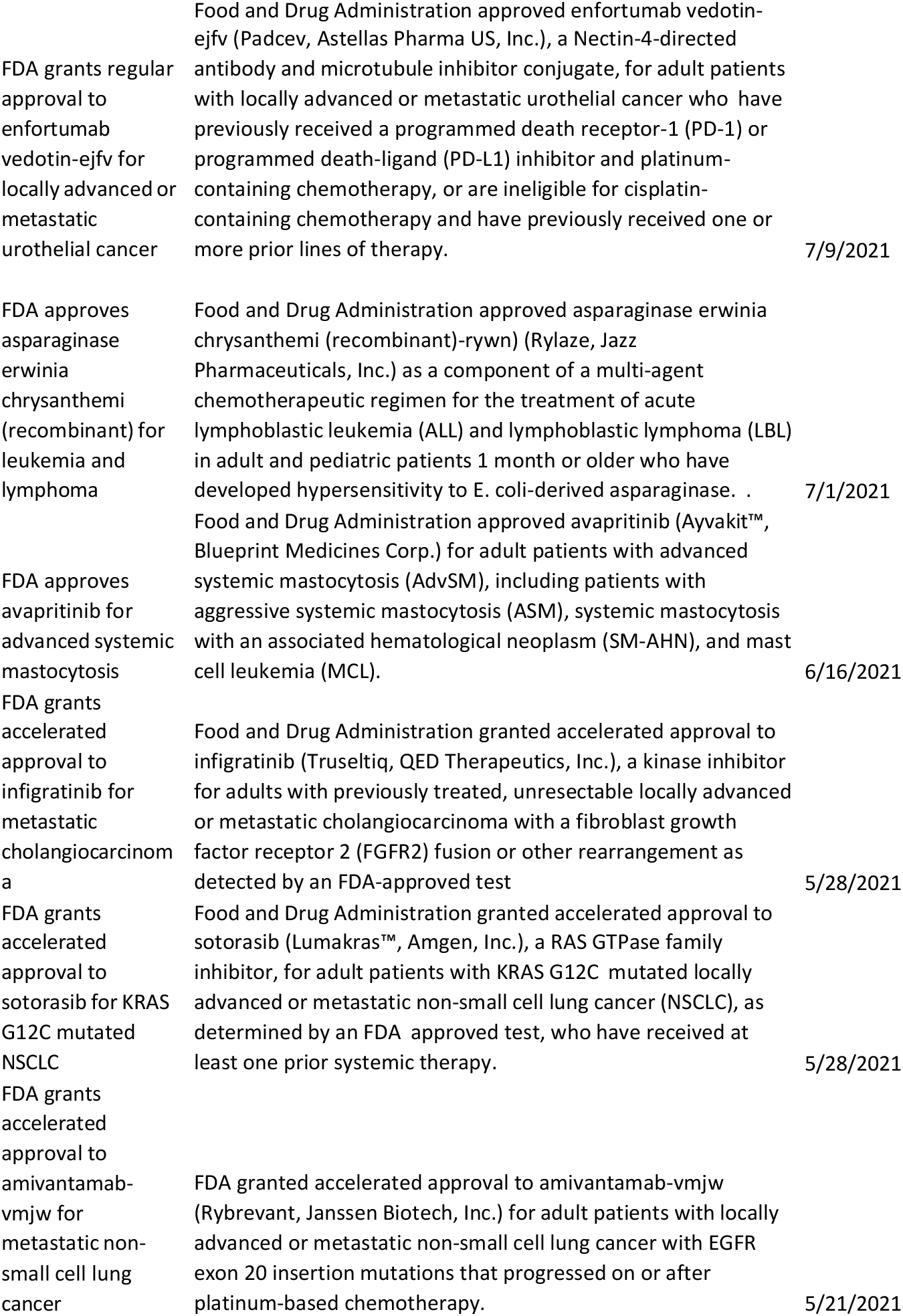
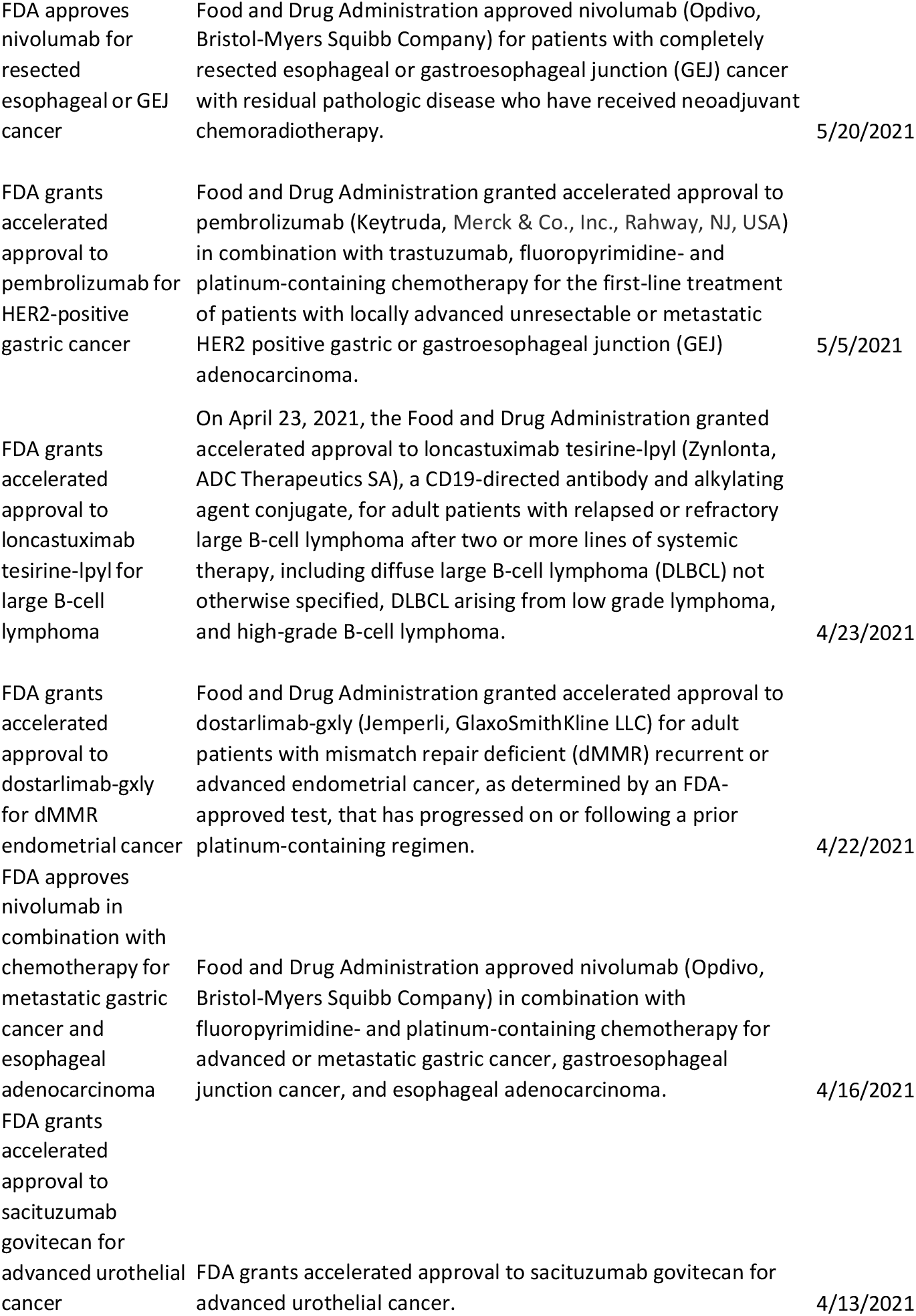
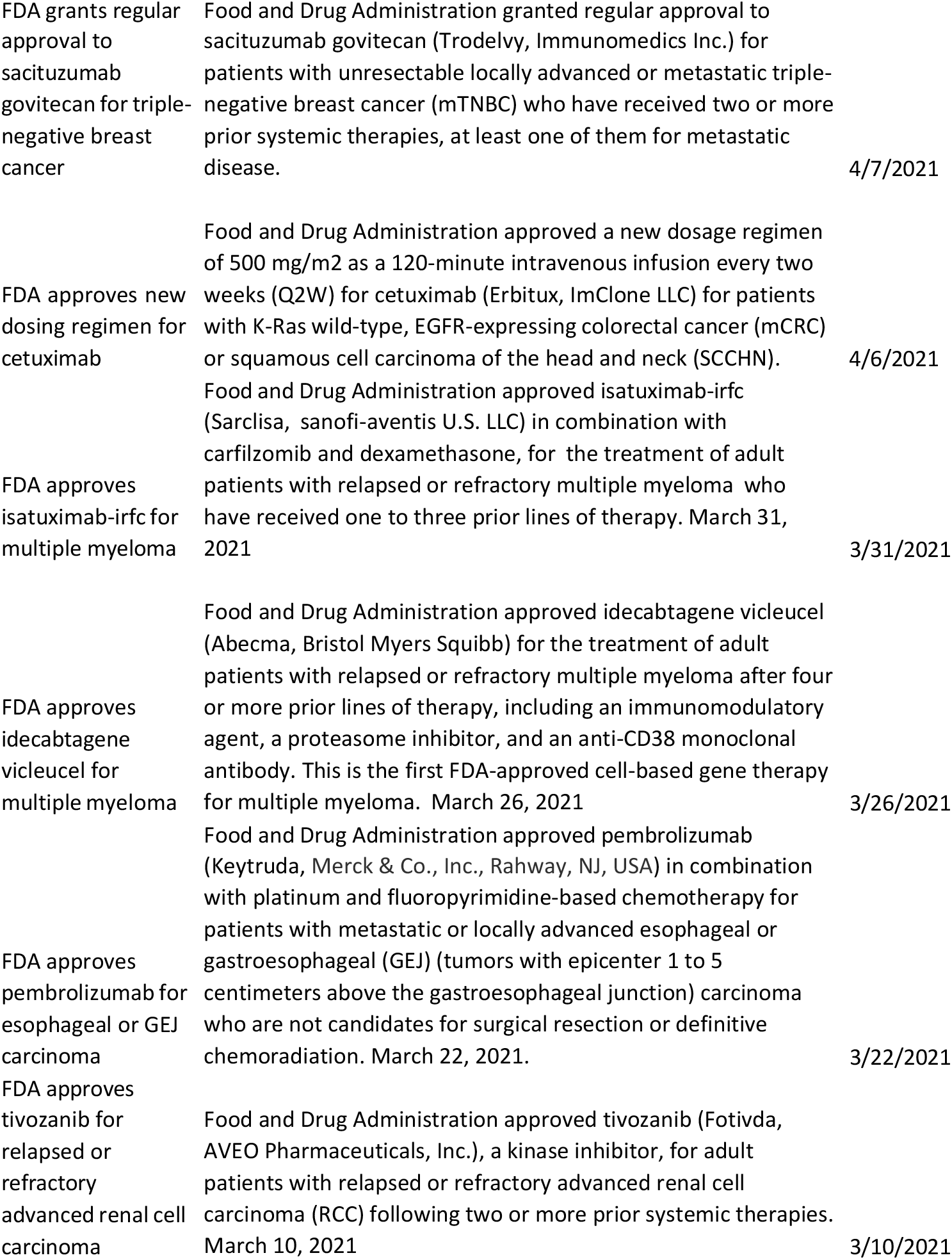
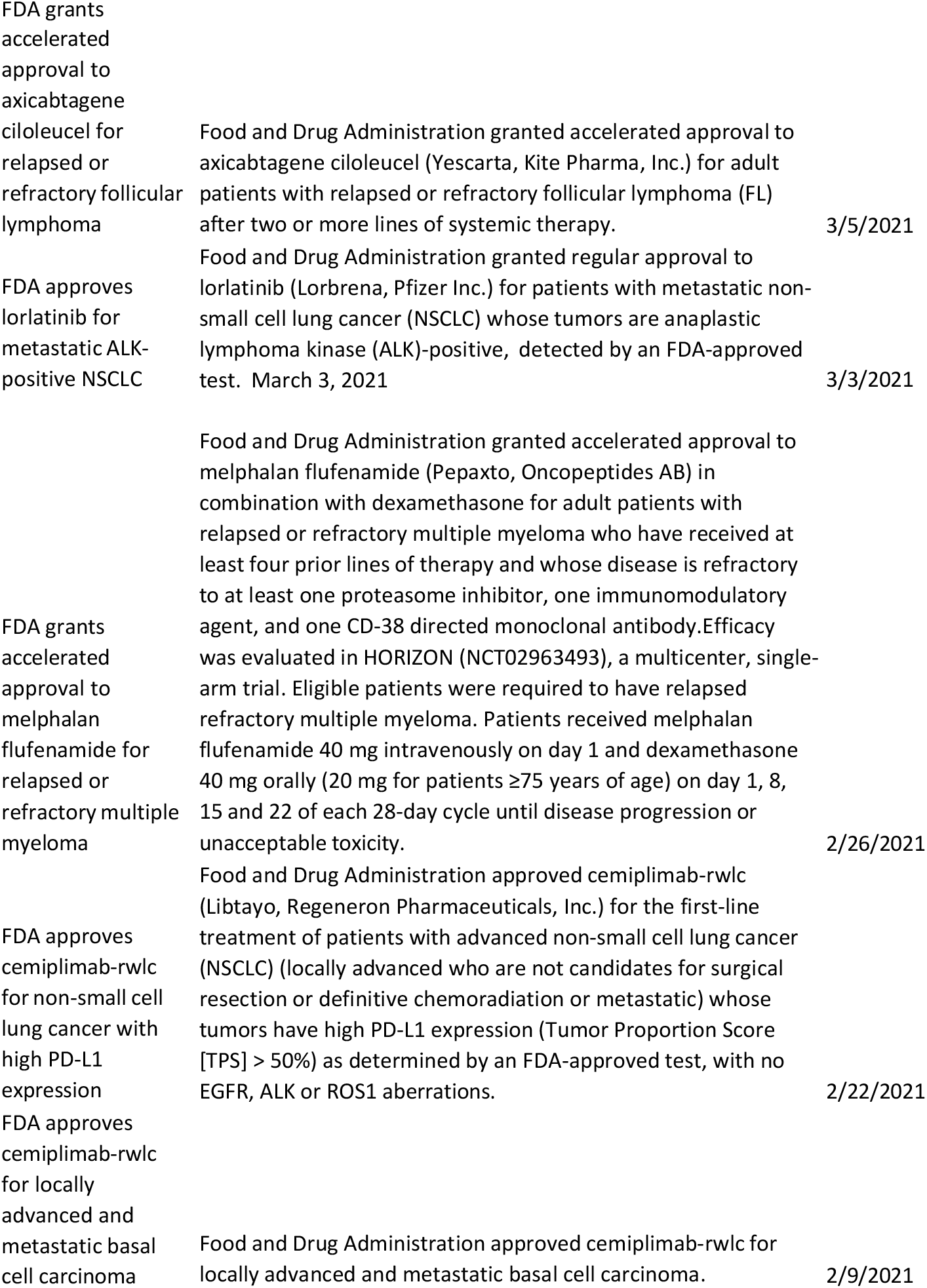
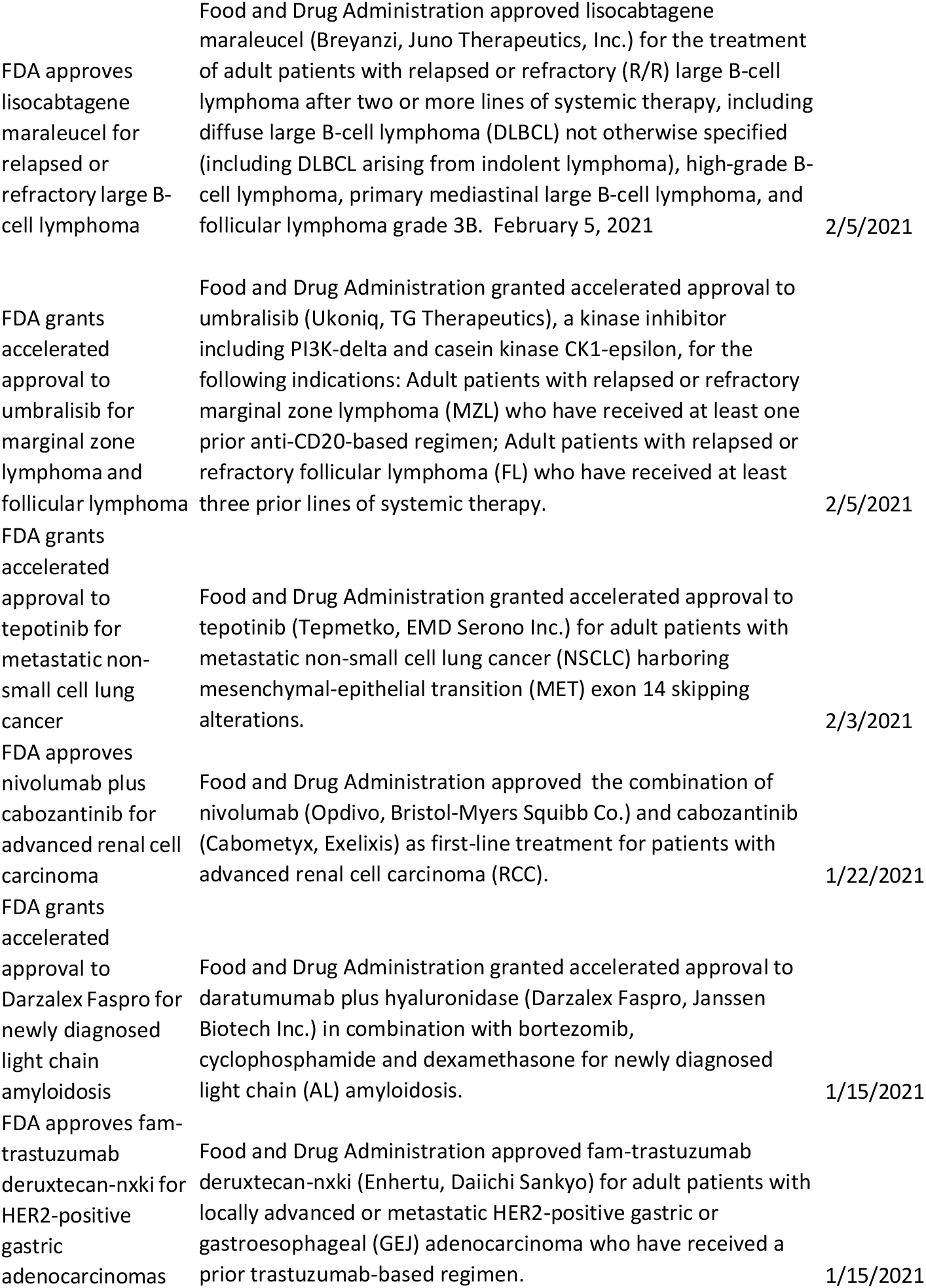
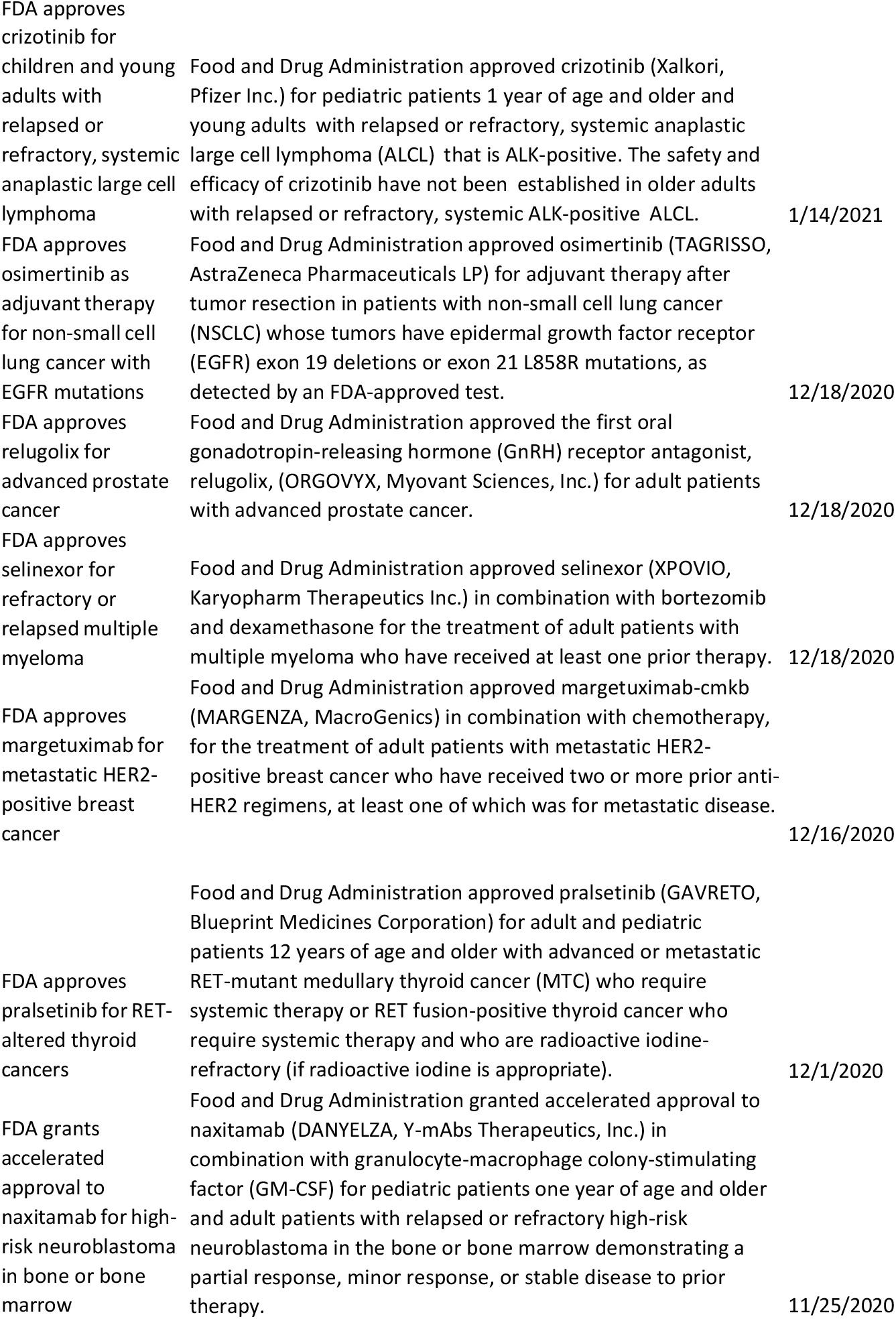
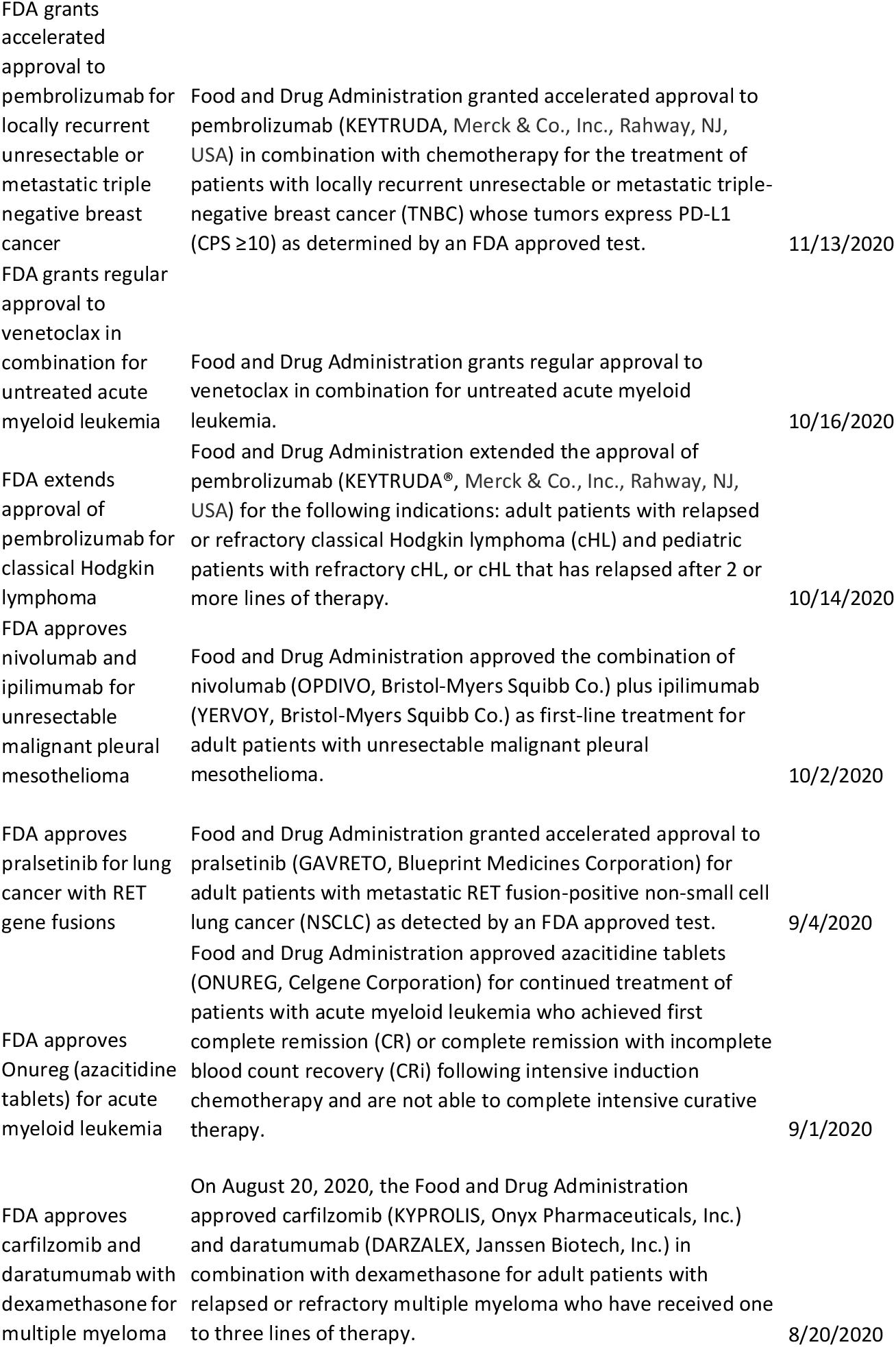
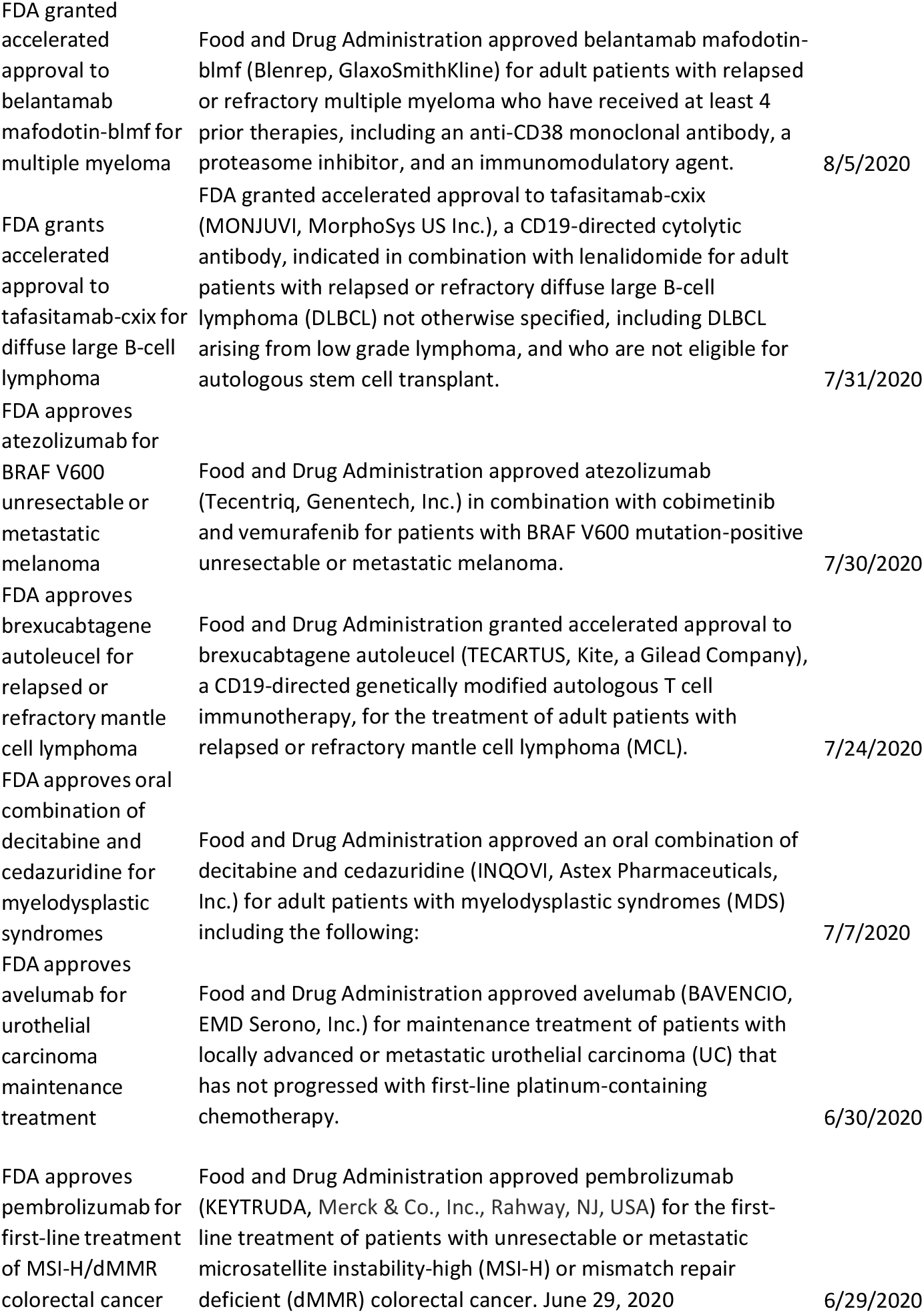
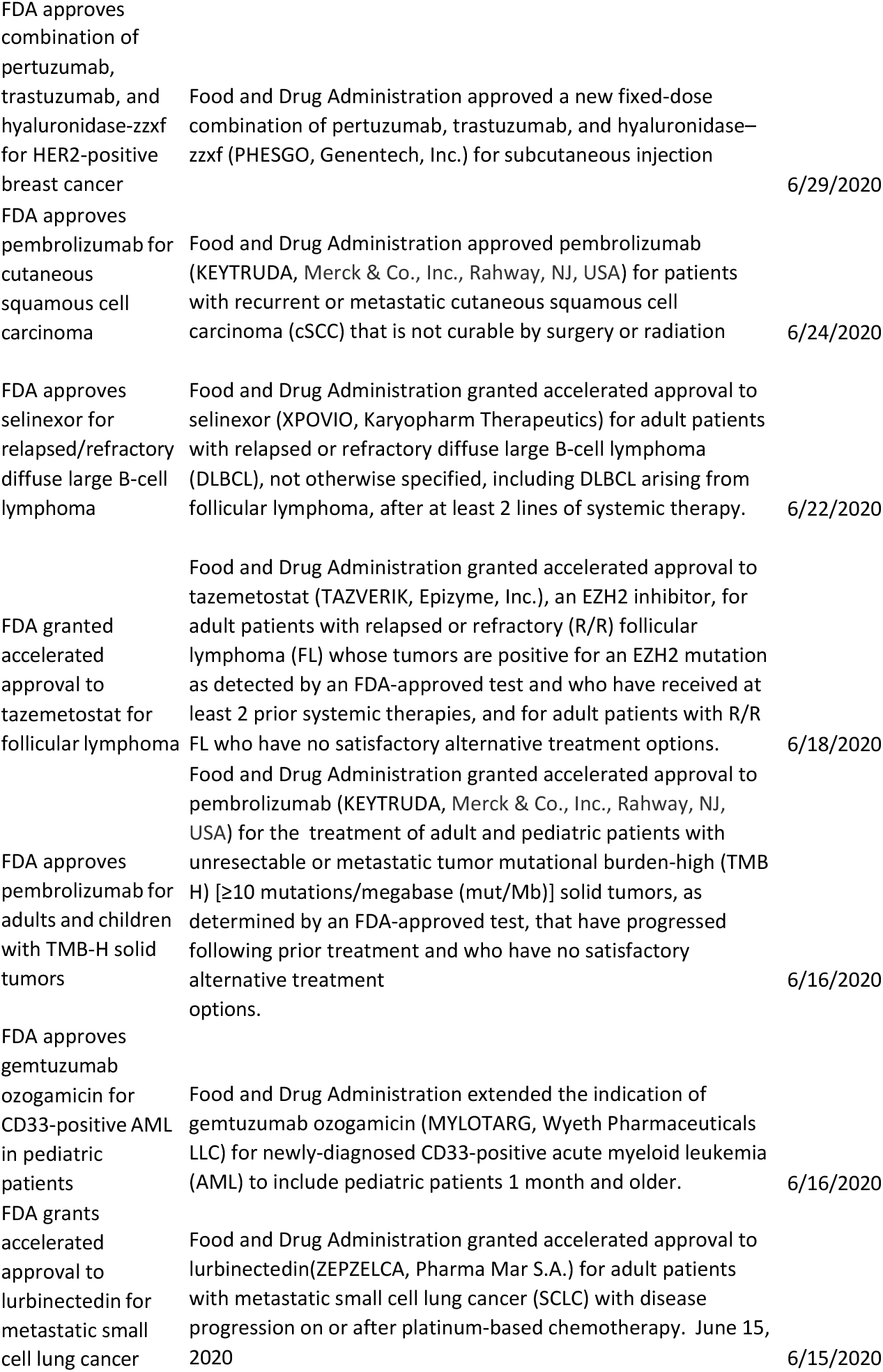
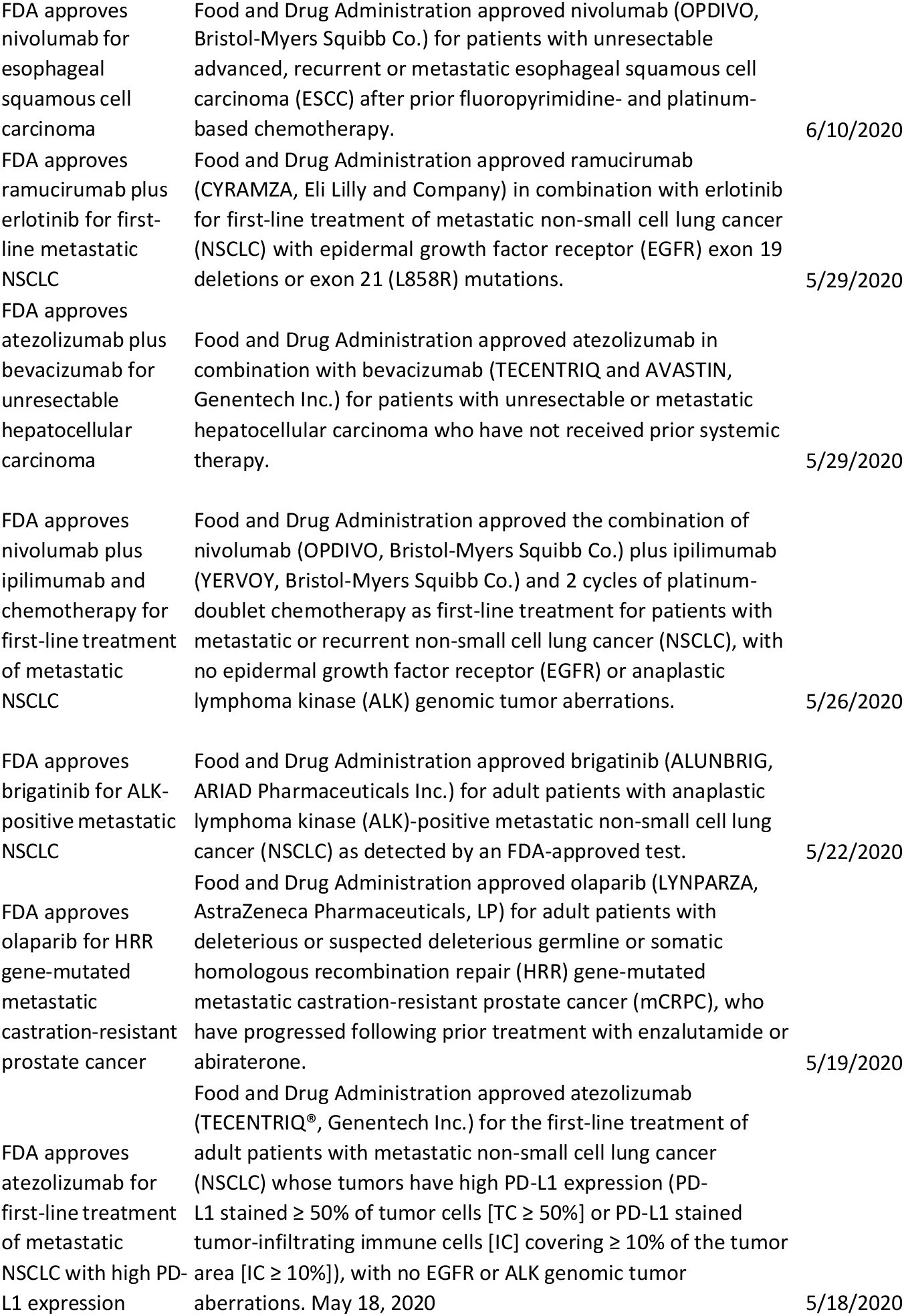
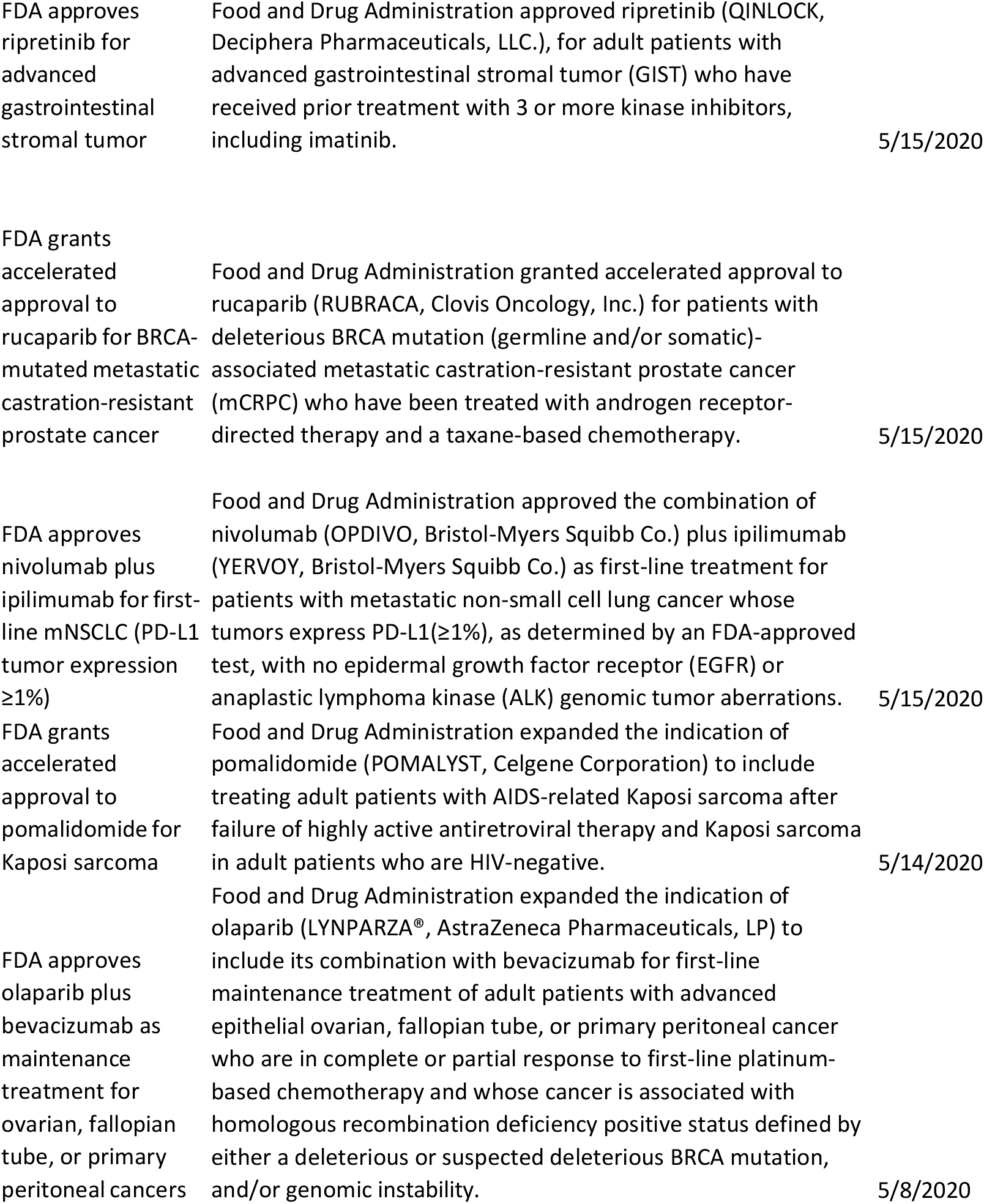
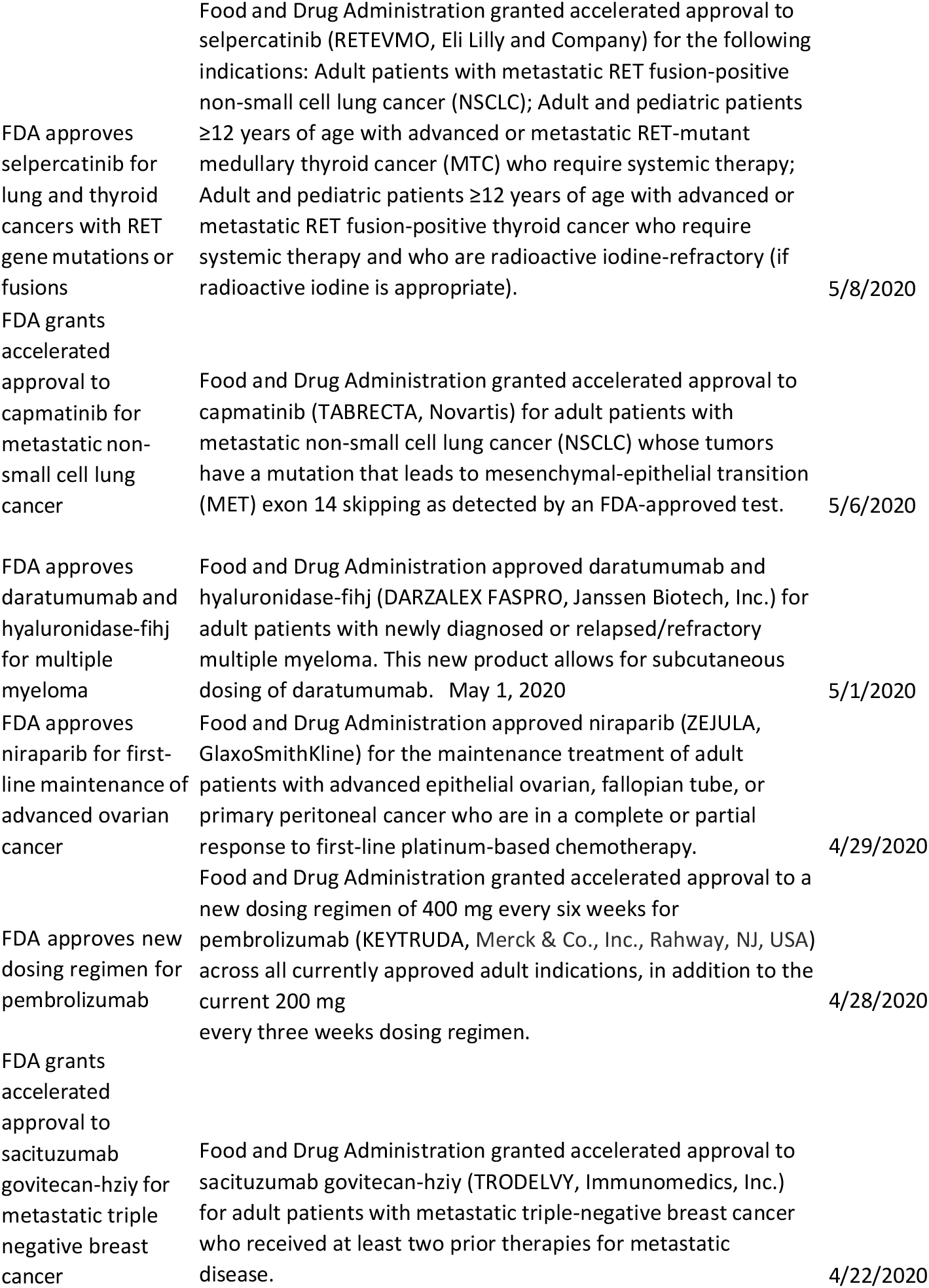
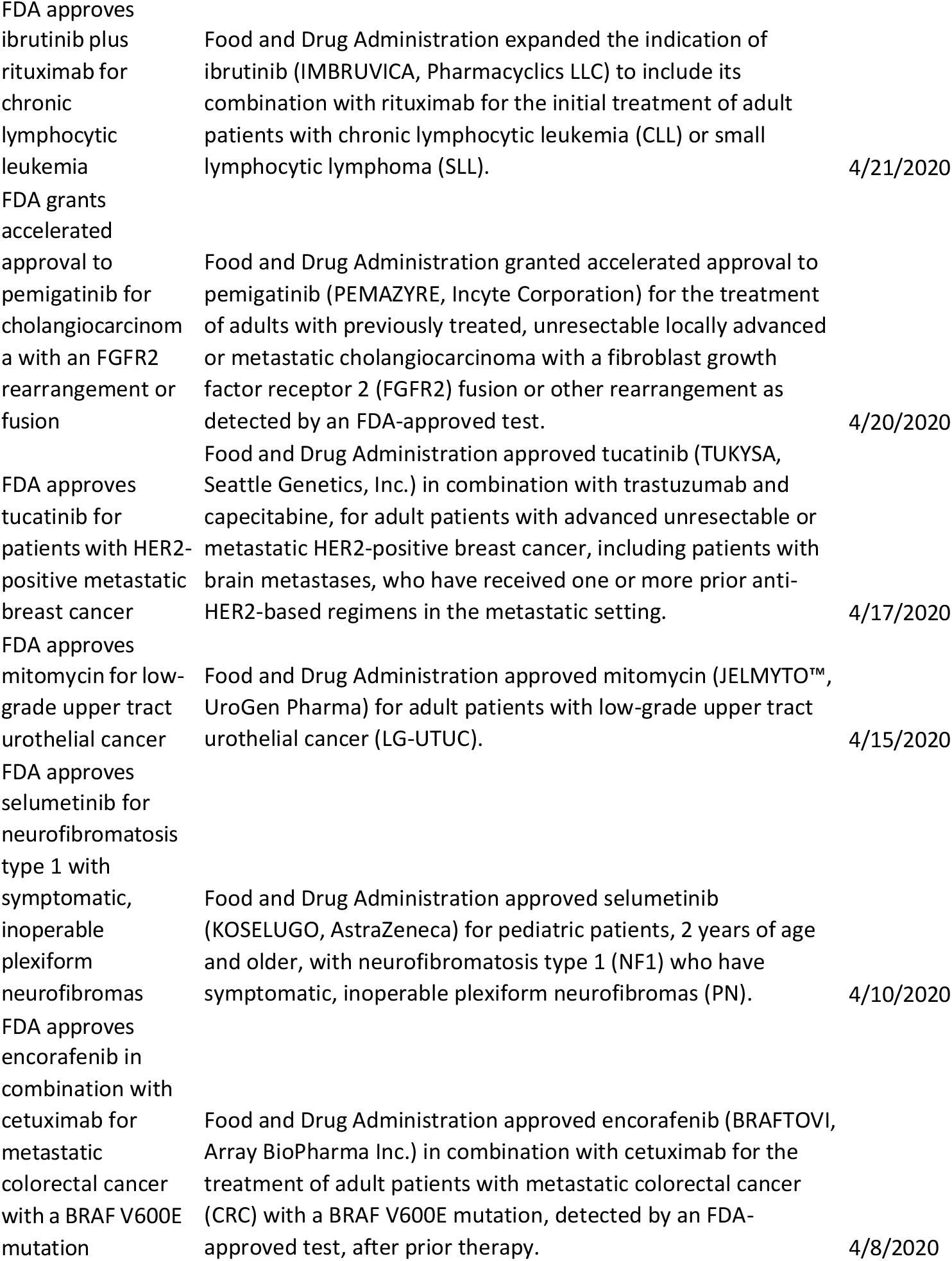
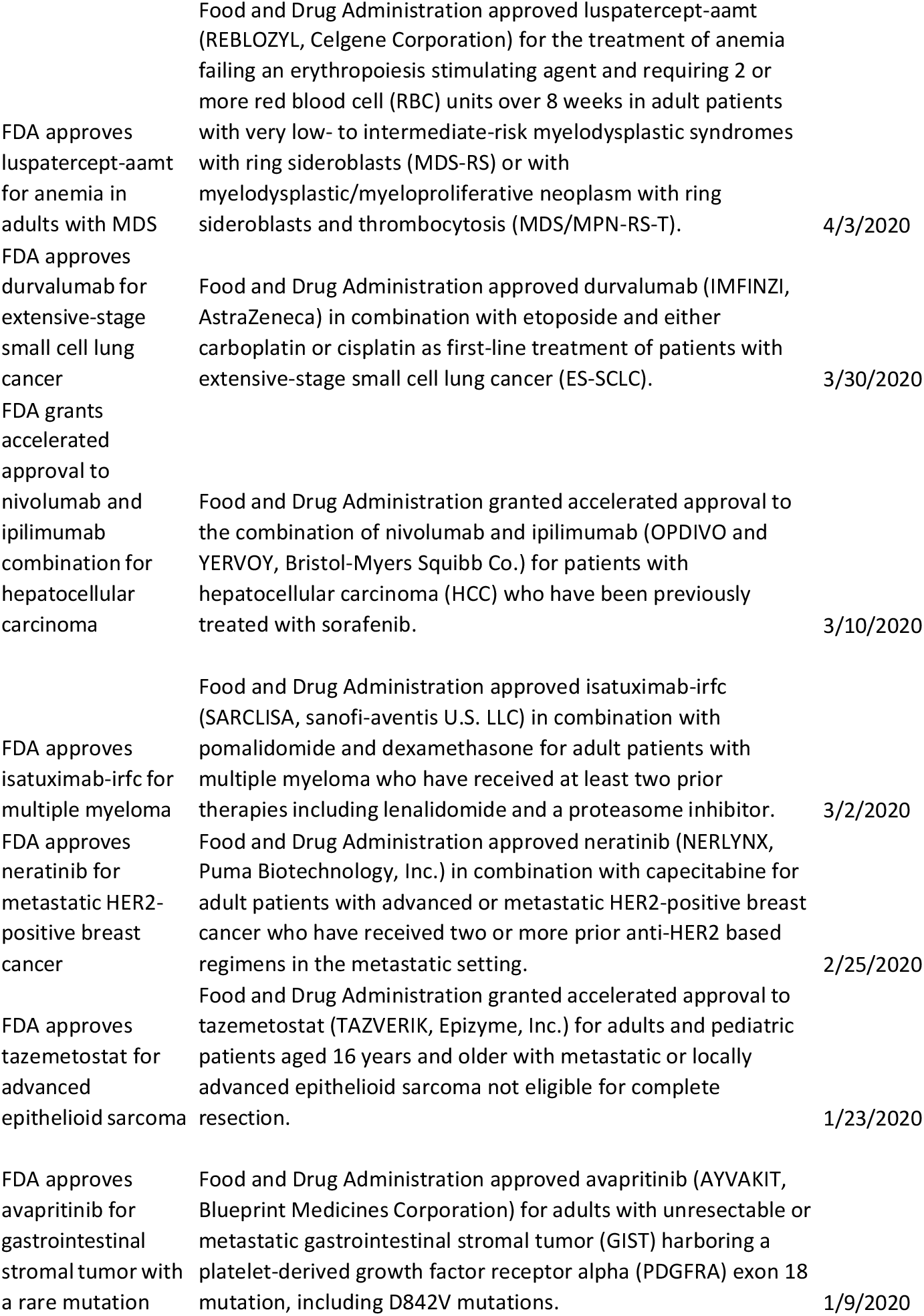
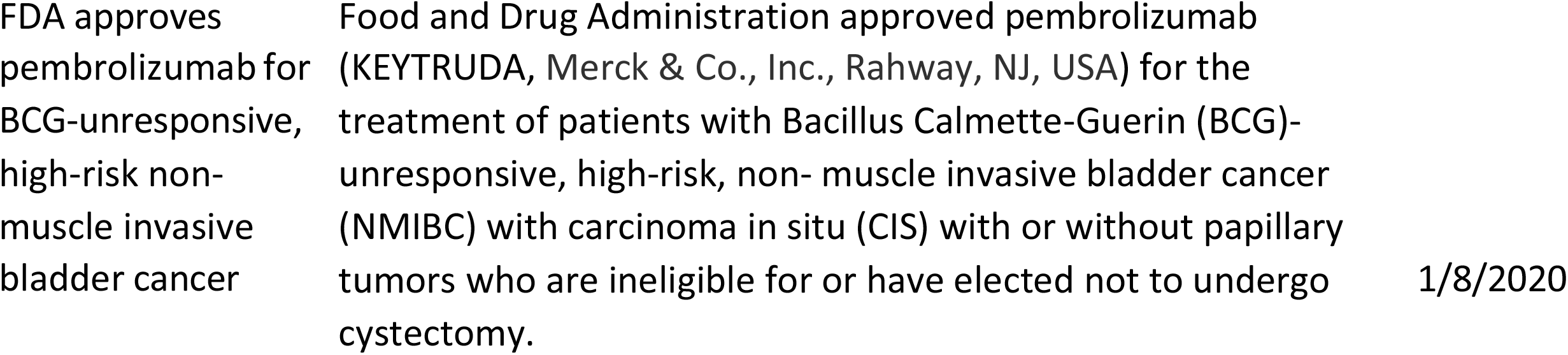
Oncology Approvals by the US FDA, 2020-2021.

